# Does Obstructive sleep apnea increase the risk of Parkinson Disease? A systematic review and meta-analysis

**DOI:** 10.1101/2020.10.22.20214882

**Authors:** Tianming Zhao, Kun Xuan, Chenyu Sun, Haixia Liu, Xin Chen, Yehuan Sun

**Affiliations:** Department of Epidemiology and Health Statistics, School of Public Health, Anhui Medical University, No. 81 Meishan Road, Hefei 230032, Anhui, China; AMITA Health Saint Joseph Hospital Chicago, 2900 N. Lake Shore Drive, Chicago, IL 60657, USA; Center for Evidence-Based Practice, Anhui Medical University, No. 81 Meishan Road, Hefei, 230032, Anhui, China

**Keywords:** Obstructive sleep apnea, Parkinson Disease, Cohort studies, Systematic review

## Abstract

**Objectives:** The evidence of whether the obstructive sleep apnea (OSA) could increase the risk of Parkinson disease (PD) remains controversial. This meta-analysis was conducted in hopes of providing evidence of associations between the OSA and the risk of PD incidence.

**Methods:** Relevant studies published until 7 September, 2020 were retrieved from 6 databases. 6 studies were finally included based on our selection criteria. Hazard Ratios (HRs) and their 95%CI of each study were collected to evaluate the relationship between obstructive sleep apnea and Parkinson disease risk. Subgroup analysis was performed based on gender and sample size.

**Results:** We find a positive association between obstructive sleep apnea and Parkinson disease risk (HR=1.64, 95%CI:1.56-1.73, I^2^=23.8%). OSA patients have a higher risk of Parkinson disease than subjects without OSA, especially in male patients (HR=1.56, 95%CI: 1.30-1.87, I^2^=0.0%). Based on results of sensitivity analysis, our study results were quite stable.

**Conclusions:** Our results provided evidence of a modest positive association between obstructive sleep apnea and Parkinson disease. It is a certain degree of significance to improve our understanding of obstructive sleep apnea and take measures to prevent and treat it. Moreover, additional studies are recommended to explore this topic in more detail. This study was registered in PROSPERO (CRD42020207528).

## Introduction

Characterized by partial or complete collapse of the upper airway during sleep several times, obstructive sleep apnea (OSA) could lead to reduced or absent airflow lasting for 10 seconds or even longer which are related to cortical arousal or a fall in blood oxygen saturation [1]. Patients with OSA may be unaware of their disease which could result in undiagnosed [2,3]. The threat of OSA to the health of general population cannot be ignored and the OSA could increase the risk of several diseases (hypertension, myocardial infarction, stroke, and diabetes, etc.) if not be treated properly [4-9]. Furthermore, two of the main pathophysiologic characteristics of OSA (intermittent hypoxia and sleep fragmentation) [10] are believed to contribute to poor health outcomes in OSA patients and the adverse effect on high oxygen-demanding organs may be more complicated [11]. Previous study has found that OSA patients showed brain abnormalities and metabolic deficits in multiple areas, and the tissue injury was presumed to be related to hypoxemia-induced processes [12]. Thus, clarifying the complications and potential health outcomes of OSA patients is of great importance both for healthcare and understanding of underlying mechanism.

Neurological conditions are the leading source of disability worldwide and as an important type of them, Parkinson’s disease (PD) has been growing rapidly in prevalence, disability, and deaths these years [13,14]. Parkinson’s disease is associated with neuronal injury and loss in the substantia nigra pars compacta, which is the major source of dopaminergic projections to basal ganglia; Parkinson’s disease has could affect sensory, emotional, cognitive, and autonomic functions [15,16]. The prevalence of Parkinson’s disease tends to increase with age [17] which is consistent with OSA [18]. These years, the potential association between sleep disorders and Parkinson disease has been widely concerned [19]. Previous studies indicated that the intermittent hypoxemia caused by OSA may increase the loss of dopaminergic neurons in brain areas related to PD, and OSA-related sleep fragmentation may also have adverse effects on the brain through decreasing neuronal excitability in regions including the locus coeruleus [20,21]. Besides, the Parkinson’s disease comorbidity with obstructive sleep apnea (OSA) is another serious health problem and it was reported that the PD patients with OSA had higher morbidity of fatty liver disease and hypertension [22]. Although the prevalence of obstructive sleep apnea may be higher in Parkinson Disease patients compared to those without PD, some studies confirmed while others refuted the relationship of OSA with some risk factors and symptoms in patients with PD [23-25], which indicated that the potential correlation between OSA and PD is still not clear enough. Thus, our meta-analysis is needed to implement. In our study, we aimed to illuminate whether obstructive sleep apnea patients have an increased risk of Parkinson Disease incidence based on several published epidemiological studies.

## Methods

We followed the Preferred reporting items for systematic reviews and meta-analysis (PRISMA) statement [26] to conduct our meta-analysis. Ethical approval was not applicable in this meta-analysis. The PICO question for our review is as follows: in human subjects, does having obstructive sleep apnea (compared to subjects without obstructive sleep apnea) increase the incidence of Parkinson’s disease? (Participants: human subjects; Indicator/Exposure: obstructive sleep apnea; Comparison: no obstructive sleep apnea; Outcomes: Parkinson’s disease) The protocol of this study was registered (PROSPERO ID: CRD42020207528; https://www.crd.york.ac.uk/prospero/display_record.php?RecordID=207528).

### Literature search

The Literature search was performed through six databases including PubMed, ScienceDirect, Web of Science, China National Knowledge Infrastructure (CNKI), Wanfang database and Chinese VIP database up to 7 September, 2020 using the following search terms: (obstructive sleep apnea OR sleep apnea OR OSA OR obstructive sleep apnea syndrome OR OSAS) AND (Parkinson’s disease OR Parkinson disease OR Parkinson OR PD). Relevant Chinese terms were applied to search the Chinese Databases. Only English and Chinese articles were included in this study.

### Eligibility criteria

Eligible studies were selected by two independent authors (T.Z. and K.X.) based on such including criteria: (1) Observational studies had a focus on whether people with obstructive sleep apnea are more susceptible to Parkinson disease based on results that reported Risk Ratio/Hazard Ratio/Odds Ratio and the related Standard Error (SE) or 95% confidence intervals (CI) or provided enough raw data to evaluate them; (2) Case-control studies or cohort studies; (3) Studies were published in Chinese or English. The exclusion criteria were as follows: (1) Reviews, case reports, editorials, meeting abstracts or other studies with interventional designs; (2) Studies lacked necessary information for our meta-analysis or used inappropriate statistical methods; (3) Studies with duplicate data published previously.

### Data extraction and quality assessment

Two investigators (H.L. and X.C.) independently extracted the following information from every included article: first author’s name, year of publication, study location, study design, age range of participants, sample size, total number of cases and controls (OSA patients or not), outcomes of interest (effect values and 95%CI, prevalence of PD in OSA patients), exposure (OSA) and outcome (PD incidence) assessment, and adjusted covariates. Any disagreement will be resolved by discussion until an agreement is reached or by consulting a third author. We utilized the Newcastle-Ottawa Quality Assessment Scale (NOS) [27,28] to assess the quality of included cohort studies and case-control studies. The NOS scale included 8 items, which could be stratified by three dimensions: selection, comparability, and outcome (for cohort studies) or exposure (for case-control studies) [29]. In our study, 7-9, 4-6 and 0-3 points represented high-quality, middle quality and low quality respectively [28].

### Statistical analysis

For each included case-control and cohort studies evaluating the association between OSA and PD incidence, we obtained the HRs with 95% CIs and transformed HR and their 95%CI to their logarithms and standard errors (SEs) for further quantitative analysis. We pooled HRs with 95%CIs to evaluate the potential link between OSA and Parkinson disease incidence. Besides, Cochran’s χ^2^-based Q-statistic and I^2^ were also conducted to test the heterogeneity of included studies. In Q test, p-value <0.05 was considered as a possibility of significant heterogeneity, and I^2^ statistics was used to estimate the degree of heterogeneity amongst the including studies; when I^2^ >50%, we define it as high heterogeneity and the random effect model is used in overall and subgroup analysis to confirm the most conservative results [30,31]. If not, the fixed-effect model was used [30]. Subgroup analysis was performed separately based on gender and sample size. To test the stability of pooled results, sensitivity analysis was also performed through removing each eligible study sequentially. All statistical analyses will be performed using Stata (version 14.0) software (Stata Corporation, College Station, TX, USA).

## Results

### Study selection and quality assessment

Based on our search strategy, a total of 758 records were found. According to our inclusion and exclusion criteria, we finally selected 6 studies published from 2015 to 2020 (five retrospective cohort studies and one prospective cohort studies) which included more than 1.1 million individuals [25, 32-36]. Among them, 5 retrospective cohort studies were utilized to perform meta-synthesis [25,32,33,35,36]. The details of selection are shown in Figure 1. Among these inclusive studies, one study was conducted in the USA, one study was conducted in Canada, and another four was conducted in Taiwan of China. The main characteristics of these studies are listed in Table 1. In the included prospective cohort study aiming to explore predictors of parkinsonism in idiopathic REM sleep behavior disorder (iRBD) [34], researchers analyzed several potential variables including sleep apnea (defined as apnea-hypopnea index cut-off≥15/h) among patients with iRBD. Considering the limited generalizability to general population, we didn’t include this study into our meta-synthesis. Table 2 shows the results of quality assessment of included studies. All study scored between 6 and 7 points. Among them, two studies are middle-quality [33,34] while rest of them are high-quality [25,32,35,36].

**Table 1.** Characteristics of included studies.

| First author | Published year | Study location | Study design | Mean age/age range | Sample size | OSA ascertainment | PD ascertainment | Effect value(95%CI) |
| --- | --- | --- | --- | --- | --- | --- | --- | --- |
| Chen [25] | 2015 | Taiwan | Retrospective cohort study | 45.6 | 29,133 | ICD-9-CM codes 780.51, 780.53 or 780.57 | NA | 1.84(1.40-2.43)<sup>a</sup> |
| Chou [32] | 2017 | Taiwan | Retrospective cohort study | ≥40 | 11,664 | ICD-9-CM codes 780.51, 780.53, 780.57 or 327.23 | Receiving any outpatient or inpatient care for PD (ICD-9-CM 332) | 1.85(1.02-3.35)<sup>b</sup> |
| Kummer [33] | 2019 | USA | Retrospective cohort study | 75.9 | 1,035,536 | NA | Separate outpatient claims for idiopathic PD (ICD-9-CM 332) | 1.65(1.56-1.75)<sup>c</sup> |
| Postuma [34] | 2019 | Canada | Prospective cohort study | 66.3 | 1,280 | AHI ≥15 | Neurologist testing | 0.92(0.70-1.23)<sup>d</sup> |
| Sheu [35] | 2015 | Taiwan | Retrospective cohort study | 46.5 | 9,192 | ICD-9-CM codes 327.23, 780.51, 780.53 or 780.57 and PSG for confirmation | NA | 2.26(1.32-3.88)<sup>e</sup> |
| Yeh [36] | 2016 | Taiwan | Retrospective cohort study | NA | 33,460 | Patients met the diagnostic criteria for that disease in at least 3 outpatient service claims with codes for OSA (ICD-9-CM code 780.51, 780.53, 780.57 or 327.23) at the department or division of Chest Medicine at any hospital or local medical clinic | Patients met the relevant diagnostic criteria for that disease in at least 3 outpatient service claims with codes for PD (ICD-9-CM code 332) | 1.37(1.12-1.68)<sup>f</sup> |

**Note:** CI: confidence interval; OSA, obstructive sleep apnea; PD, Parkinson Disease; NA: not available; AHI: apnea-hypopnea index.
a, hazard ratio (HR), adjusted factors: age, sex, urbanization level, income, and comorbidity;
b, hazard ratio (HR), adjusted factors: sex, age, urbanization level, geographic region, monthly income, hypertension, hyperlipidemia, diabetes mellitus, asthma, COPD, and head injury;
c, hazard ratio (HR), adjusted factors: age, sex, race/ethnicity, and baseline Charlson comorbidities;
d, hazard ratio (HR), adjusted factors: adjusted for age, sex and recruiting center;
e, hazard ratio (HR) stratified by sex, age group and year of index ambulatory care visit and adjusted for patients' geographical location, urbanization level, and monthly income;
f, hazard ratio (HR), adjusted factors: age, sex, diabetes, hypertension, coronary artery disease, stroke, hyperlipidemia, chronic kidney disease, and gout.

**Table 2.** Newcastle - Ottawa quality assessment scale (cohort study)

| Study | Year | Selection |  |  |  | Comparability | Outcome |  |  | Total |
| --- | --- | --- | --- | --- | --- | --- | --- | --- | --- | --- |
|  |  | A | B | C | D | E | F | G | H |  |
| Chen [25] | 2015 | 1 | 1 | 1 | 1 | 1 | 1 | 1 | 0 | 7 |
| Chou [32] | 2017 | 1 | 1 | 1 | 1 | 1 | 1 | 1 | 0 | 7 |
| Kummer [33] | 2019 | 0 | 1 | 1 | 1 | 1 | 1 | 1 | 0 | 6 |
| Postuma [34] | 2019 | 0 | 1 | 1 | 1 | 1 | 1 | 1 | 0 | 6 |
| Sheu [35] | 2015 | 1 | 1 | 1 | 1 | 1 | 1 | 1 | 0 | 7 |
| Yeh [36] | 2015 | 1 | 1 | 1 | 1 | 1 | 1 | 1 | 0 | 7 |

A: Representativeness of the exposed cohort; B: Selection of the nonexposed cohort; C: Ascertainment of exposure; D: Demonstration that outcome of interest was not present at start of study; E: Comparability of cohorts on the basis of the design or analysis; F: Assessment of outcome; G: Was follow-up long enough for outcomes to occur; H: Adequacy of follow up of cohorts

**Figure 1.**
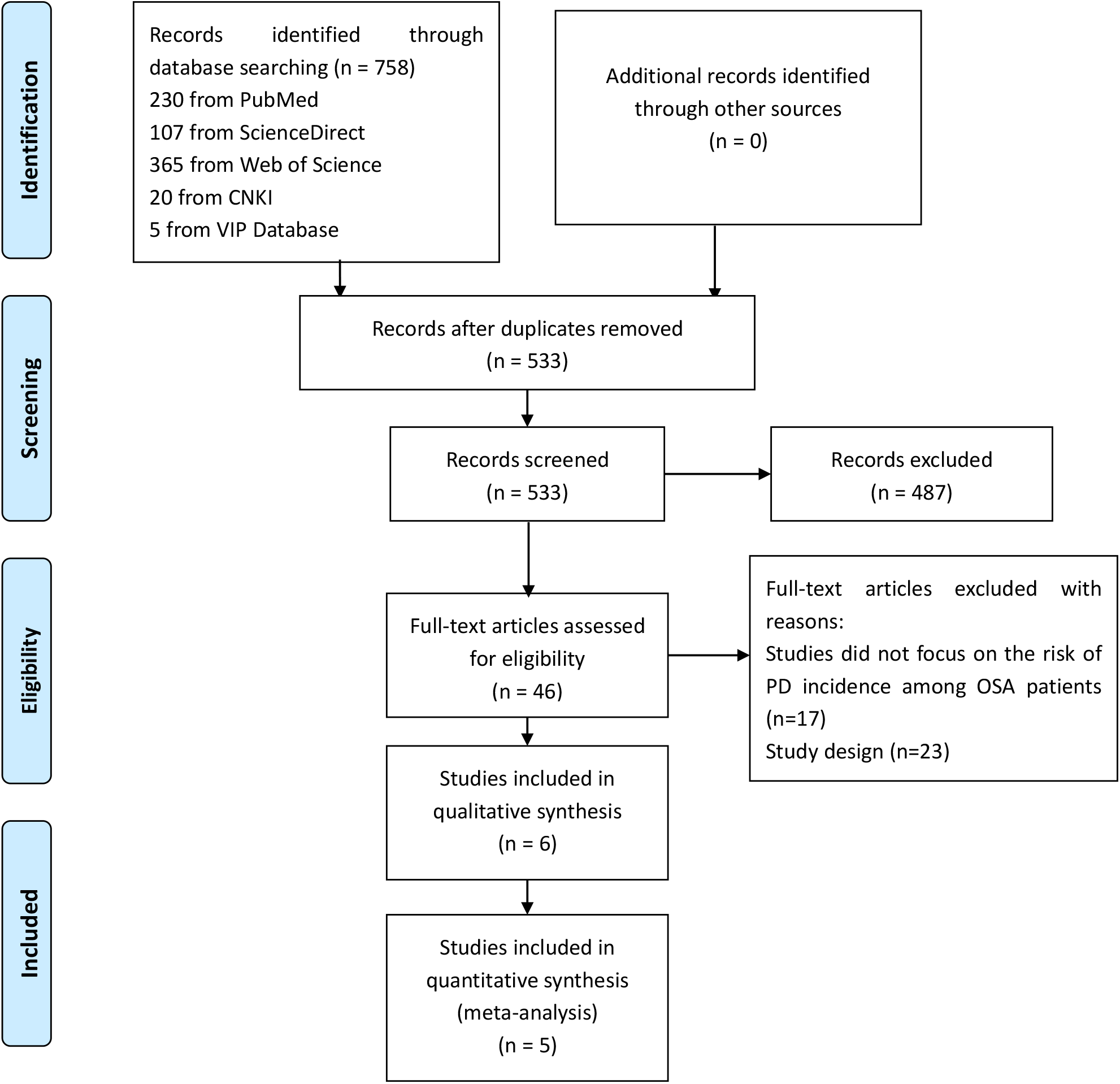
Flow chart of selecting eligible studies.

### Meta-synthesis of results

As shown in Figure 2.1, in our study, we found a positive relationship between the OSA and PD risk, the OSA patients have an increased risk of PD incidence based on the fixed effect model (HR=1.64, 95%CI:1.56-1.73, I^2^=23.8%).

**Figure 2.**
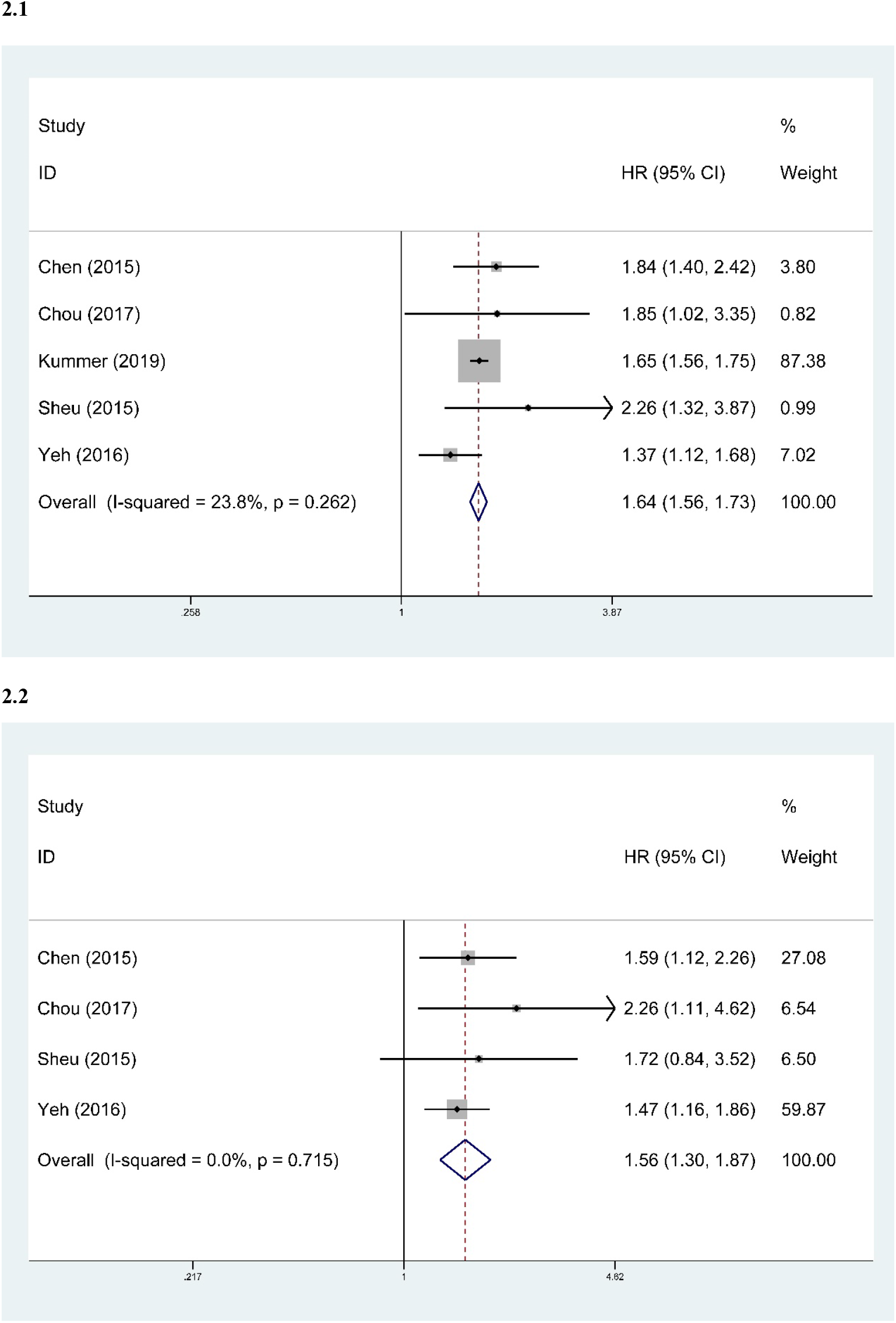
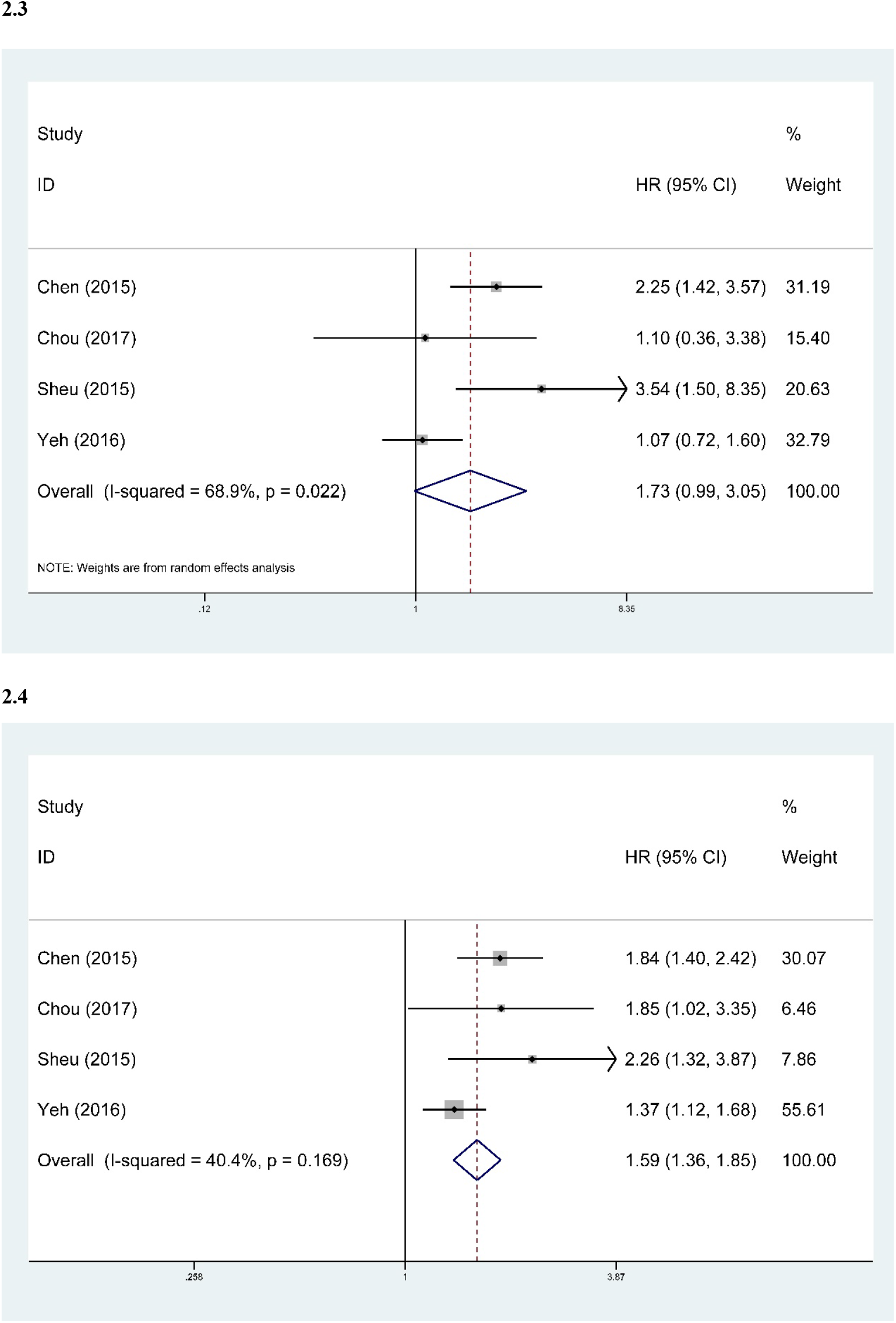
Forest plots of hazard ratio of PD risk of OSA patients; 2.1, the overall results of association between OSA and PD incidence; 2.2, the association between OSA and PD incidence among male patients; 2.3, association between OSA and PD incidence in female patients; 2.4 the association between OSA and PD incidence in studies with relatively small samples.

Subgroup analysis was conducted based on gender and sample size. In gender, male patients with OSA have an increased risk of PD incidence (HR=1.56, 95%CI:1.30-1.87, I^2^=0.0%) (Figure 2.2). However, in female patients, the association was not significantly positive based on random effect model (HR= 1.73, 95%CI:0.99-3.05, I^2^=68.9%) (Figure 2.3). As for sample size, we omitted study with the largest sample [33] and synthesize other studies, we found positive association between the OSA and PD in studies with relatively small samples (HR=1.59, 95%CI:1.36-1.85, I^2^=40.4%) (Figure 2.4).

#### Sensitivity and publication bias analysis

We applied Egger’s test and Begg’s test to detect publication bias. Based on Begg’s test (p=0.806) and Egger’s test (p=0.728), the publication bias was acceptable and there was no evidence of distinct publication bias. According to the results of sensitivity analysis, the resulted was not changed when omitting each study sequentially, which indicated that our study results were quite stable.

## Discussion

In our study, we found that obstructive sleep apnea patients have a higher risk of PD incidence (about 1.64 times higher than that in subjects without OSA) which synthesize studies covering areas from Asia and North America. According to subgroup analysis, when it comes to male patients, the correlation was still positive (the risk of PD in male OSA patients was 1.56 times higher than male subjects without OSA). Considering the high heterogeneity in female patients (I2=68.9%), we performed random effect model to evaluate the correlation in female patients. The risk of PD in female OSA patients was 1.73 times higher than female subjects without OSA though didn’t significantly correlated. Besides, the results of subgroup analysis also indicated that gender may be the main source of heterogeneity. The potential reasons may as follow: firstly, limited by the sample size of female patients with or without OSA, our study results of female were not stable enough. Among individuals in subgroup analysis regarding to gender, female subject only accounted for approximately 27.8%, which indicated that future studies are needed to explore the correlation in female patients based on a larger sample to get more stable results. Secondly, previous studies have demonstrated that OSA is more frequent in men than women with a male-to-female ratio 3:1 in the general population [37-39], which could affect the female OSA patients’ enrollment and lead to the difference of sample size between gender.

As for the underlying mechanisms of PD risk among OSA patients, previous studies proposed that the main characteristics of OSA (intermittent hypoxia and sleep fragmentation) could lead to structural and functional changes in brain, which were closely related to a series of neurodegenerative disorders including PD [40,41]. To be specific, OSA-related intermittent hypoxemia has been proven to increase numerous products of oxidation, notably reactive oxygen, and nitrogen species, which could cause cognitive decline in the elderly, as well as in neurodegenerative diseases by injuries from oxidative stress [40,42,43]. It is also proposed that intermittent hypoxemia has been linked to dysfunction of the blood–brain barrier, which is critical to maintain brain homeostasis [40]. Besides, previous study has also implied that sleep fragmentation may contribute to the development of PD pathology through promotion of oxidative stress or impairment of the clearance of toxic proteins [44]. After all, OSA-related oxidative stress and inflammation have been believed to be involved in the pathophysiology of PD [36,45,46]. However, in the prospective cohort study of Postuma et al. [34], sleep apnea was not related to parkinsonism among subjects with iRBD (HR= 0.92, 95%CI: 0.7-1.2). Although the generalizability of subjects included in this study to general population was limited, the results of this work have also indicated that the underlying mechanism of PD risk among OSA patients is not clear enough and still needed to be explored by experimental studies in the future.

Compared to previous meta-analysis [47], our current meta-analysis has several strengths. First, our study further confirmed the association between OSA and PD incidence in a larger sample and covered a wider range of people including Asia and North America. We also used different methods to synthesize HRs in female patients. We performed random-effect model in order to get more conservative results and discussed the difference results of female subjects. Limited by the participants included in our study, the association was not stable enough and future studies still need to explore the potential correlations in female patients with larger sample size and high-quality study design. Besides, Sun et al. [47] only analyzed gender in their subgroup to explore the correlations between OSA and PD, in our study, we performed gender and sample size in our subgroup analysis in order to produce more convincing evidence.

There are several limitations in our meta-analysis. First, according to our study, the gender difference may be the main source of heterogeneity existing heterogeneity among the included studies, which makes our results needed to be improved and enriched in the future. Secondly, limited number of our included studies and the study design (retrospective cohort study) makes our study results needed to be further confirmed by more high-quality studies (prospective cohort) in the future. Thirdly, participants included in our studies merely comes from Taiwan of China and the USA, which limited the generalizability to general population in other countries.

## Conclusion

In conclusion, our meta-analysis further confirmed that obstructive sleep apnea is significantly associated with an increased risk of Parkinson disease especially among male individuals. Future high-quality studies are needed to confirm our study results in female population and mechanisms of this potential relationship are needed to be explored as well.

## Data Availability

Some or all data, models, or code generated or used during the study are available from the corresponding author by request.

